# Heart Rate Variability in Patients with Cirrhosis: A Systematic Review and Meta-analysis

**DOI:** 10.1101/2021.01.20.21249506

**Authors:** Tope Oyelade, Gabriele Canciani, Gabriele Carbone, Jaber Alqahtani, Kevin Moore, Ali R. Mani

## Abstract

**Background:** Cirrhosis is associated with abnormal autonomic function and regulation of cardiac rhythm. Measurement of heart rate variability (HRV) provides an accurate and non-invasive measurement of autonomic function as well as liver disease severity currently calculated using the MELD, UKELD, or ChildPugh scores. This review assesses the methods employed for the measurement of HRV, and evaluates the alteration of HRV indices in cirrhosis, as well as their value in prognosis.

**Method:** We undertook a systematic review using Medline, Embase and Pubmed databases in July 2020. Data were extracted using the Preferred Reporting Items for Systematic Reviews and Meta-Analyses (PRISMA) guidelines. Risk of bias of included studies was assessed by a modified version of the Newcastle Ottawa Scale. The studies descriptive were analysed and the standardized mean differences of HRV indices were pooled.

**Results:** Of the 247 studies generated from our search, 14 studies were included. One of the 14 studies was excluded from meta-analysis because it reported only median of HRV indices. The studies included have a low risk of bias, and include 583 patients with cirrhosis and 349 healthy controls. The HRV time and frequency domains were significantly lower in cirrhotic patients. Between-studies heterogeneity was high in most of the pooled studies (P<0.05). Further, HRV indices predict survival independent of the severity of liver disease as assessed by MELD.

**Conclusion:** HRV is decreased in patients with cirrhosis compared with healthy matched controls. HRV correlated with severity of liver disease and independently predicted survival. There was considerable variation in the methods used for HRV analysis, and this impedes interpretation and clinical applicability. Based on the data analysed, SDNN (standard deviation of inter-beat intervals) and cSDNN (i.e. SDNN corrected for basal heart rate) are the most suitable indices for prognosis in patients with cirrhosis.

## Introduction

Liver cirrhosis accounts for more than one million deaths annually worldwide, with numbers increasing year on year (1-3). However, patients with cirrhosis have a range of conditions from early uncomplicated cirrhosis which is asymptomatic, to decompensated cirrhosis where organ system start to fail and patients present with many complications such as ascites, hepatic encephalopathy and variceal bleeding (4, 5). Once a patient starts to develop complications, various scoring systems including MELD, MELD-Na, or UKELD are used to calculate prognosis and the need for liver transplant at the bedside or in the clinic. These scoring systems are widely available using Apps on smartphones or web-based calculators. However, the scoring systems do not take into account the alteration in autonomic nervous system (ANS) observed in cirrhosis (6).

A simple and very useful tool to assess the state of the ANS is heart rate variability (HRV). HRV is the variation over time of the intervals between consecutive normal heartbeats (NN). Physiologically, instantaneous heart rate variation represents the capacity to adapt the heart rate (HR) to different internal and environmental circumstances and is modulated by the ANS (7). Tsuji et al. for the first time demonstrated the prognostic value of HRV analysis in the Framingham cohort study and reported that individuals with reduced HRV had increased risk for all-cause mortality (8). HRV provides a non-invasive evaluation of autonomic regulation of the cardiac rhythm and indexes the interplay between the intrinsic cardiac rhythm and external regulatory controls (9). Importantly, with medical advances, it is becoming increasingly recognized that calculation of HRV from a continuous ECG tracing provides additional and clinically useful information to clinicians on both the severity and prognosis of patients with cirrhosis (10).

Although autonomic dysfunction and cirrhotic cardiomyopathy are well established complications of cirrhosis (11, 12), these are not assessed by MELD, UKELD or Child-Pugh. This is particularly important since recent reports suggest that HRV predicts survival in cirrhosis independently of MELD and Child-Pugh and therefore may provide additive information currently lacking in existing scoring systems (13-15). However, for HRV to be of value in research and clinical practice, there needs to be standardization of processes that enables simple and accurate assessment of HRV. This includes standardization of the ECG recording techniques, duration of recording, methods and clinical interpretation of HRV and also the availability of these in the point of care via mobile or web-based apps. The aim of this study is to analyse the methods used to record and report HRV in the literature, assess HRV difference between patients with cirrhosis and controls, and finally to propose the standardization of HRV measurement technique to improve interpretation and clinical and research applicability.

## Method

This systematic review follows the guideline of the Preferred Reporting in Systematic Reviews and MetaAnalysis (PRISMA) (16). Embase, Medline and Pubmed databases were searched on the 8^th^ of July 2020. An extensive search strategy using Medical Subject Headings (MeSH) terms was performed (Figure S1). Studies retrieved from the search were uploaded to Endnote and duplicates were removed. The duplicate-free studies were then uploaded to Rayyan software and title/abstract screening performed by two independent reviewers.

Furthermore, a mini-systematic review was performed to assess the use of HRV indices as prognostic markers especially in the prediction of survival in patients with cirrhosis. Included papers were simultaneously screened and references searched for studies that performed survival analysis.

### Inclusion and exclusion criteria

Only observational studies were included in the main review. Studies were considered eligible if: either of the HRV time, frequency and non-linear indices were used in assessing autonomic cardiac control in cirrhosis. We excluded studies that did not include control group; studies involving non-cirrhotic liver disease; studies involving pharmacological or non-pharmacological interventions known to affect HRV indices; and studies involving orthostatic tilting as the sole method of HRV assessment. For the mini-systematic review, studies were included if HRV indices of survivals and non-surviving patients were statistically compared irrespective of whether risk analysis was performed.

### Data collection

Titles and abstracts were independently screened for potentially eligible studies and conflict about eligibility resolved through virtual meetings. The eligible articles were then analysed to identify studies meeting the inclusion criteria. All conflicts were resolved by a third reviewer.

Based on predefined criteria, the following data were extracted: the aims; summary of findings; sample and groups size; study setting and country; etiologies of liver disease; Duration and equipment used for ECG recording; Methods of HRV analysis including data cleaning, analysed length, analysis software and indices calculated; The etiology of cirrhosis including alcohol, fatty, primary biliary cholangitis, viral and cryptogenic were also extracted (Tables 1 and 2).

**Table 1.** General characteristics of included studies.

| Author | Aim | Conclusion | Country | Setting | Liver Disease (Male) | Control (male) | Aetiology | Child-Pugh /Histological Classification |
| --- | --- | --- | --- | --- | --- | --- | --- | --- |
| **Ates F. et al. 2006 (22)** | To assess using HRV autonomic dysfunction and its correlation with severity and 2-year survival in cirrhotic patients. | (1) HRV time-domain indices significantly reduced in cirrhotic patients compared to healthy subjects. (2) HRV indices also significantly reduced in non-survivor vs survivors after 2 years of follow-up. | Turkey | NS | 30(19) | 28(16) | HBV = 22 HCV = 8 | A = 5<br>B = 11<br>C = 14 |
| **Baratta L. et al. 2010 (23)** | To assess the effect on liver transplantation (LT)) on autonomic control of cardiac function in cirrhotic patient using HRV. | (1) HRV indices (SDNN and RMSSD) was reduced in cirrhotic patients compared with healthy subjects. (2) LT corrected the reduced SDNN but RMSSD and LF/HF remained unchanged. | Italy | outpatient | 30(20) | 27(14) | HBV = 4<br>HCV = 14<br>Other (NASH, Ethanol, Mixed) = 12 | A = 5<br>B = 18<br>C = 7 |
| **Coelho, L. et al. 2001 (24)** | To evaluate autonomic function in patient with liver cirrhosis using HRV and to evaluate the relationship with severity | (1) HRV is significantly reduced in chronic liver disease. (2) Reduced HRV is correlated with severity of liver disease. (3) Autonomic dysfunction is not related to the aetiology of liver disease. (4) Markers of hepatocellular dysfunction are more accurate predictor of autonomic dysfunction compared to markers of cholestasis where SDNN correlated significantly with prothrombin (r=0.64, p=0.001) and serum albumin (r=0.40, p=0.05) but not total bilirubin. | Portugal | NS | 22 (11) | 20 | Alcohol = 12<br>HBV + HCV = 6<br>Autoimmune = 2<br>Others = 2 | A = 6<br>B = 9<br>C = 7 |
| **Frokjaer V. G. et al. 2006 (25)** | To determine if autonomic dysfunction is related to cerebral blood flow autoregulation in cirrhotic patients. | (1) Cerebral autoregulation of blood flow impaired in severe cases of liver cirrhosis. (2) Impairment of cerebral autoregulation correlated with autonomic dysfunction (3) severity of cirrhosis correlated with degree of autonomic dysfunction (4) loss of sympathetic innervation of cerebral vessels possibly linked with cerebral autoregulation dysfunction in cirrhotic patients. | Denmark | outpatients | 14 (9) | 11 (5) | Alcohol = 8<br>PBC = 4<br>Cryptogenic = 2 | A = 5<br>B = 6<br>C = 3 |
| **Iga A. et al. 2003 (26)** | To assess autonomic abnormalities in patient with Liver cirrhosis using <sup>123</sup>I-metaiodobenzylguanidine (MIBG) myocardial scintigraphy and HRV. | Autonomic dysfunction is present in cirrhotic patients and can be assessed by MIBG myocardial scintigraphy and HRV. | Japan | NS | 50 (27) | 50 (33) | HBV = 4<br>HCV = 40<br>HBV + HCV = 2<br>PBC = 4 | A = 20<br>B = 12<br>C = 18 |
| **Ko F.Y. et al. 2013 (27)** | To assess the relationship between psychological distress and clinical presentations of cirrhosis using biochemical and physiological (HRV) markers. | (1) Psychological distress was correlated with increased serum aspartate aminotransferase (AST), and reduced autonomic control of the heart (HRV). (2) Inflammation may have a role to play in cirrhotic psychological distress. | China | outpatients | 125(73) | 55(29) | Alcohol = 14<br>HBV + HCV = 83<br>Autoimmune = 5<br>Parasites = 5<br>Others = 18 | A = 43<br>B = 58<br>C = 24 |
| **Lazzeri C. et al. 1997 (28)** | To assess autonomic neuropathy in patients with non-alcoholic cirrhosis with ascites. | Ascitic patients with non-alcoholic cirrhosis have autonomic neuropathy compared with healthy controls and this is associated with significant difference in HRV indices between the groups. | Italy | outpatients | 12 (7) | 12 | HBV = 2<br>HCV = 8<br>Cryptogenic = 2 | A = 0<br>B = 5<br>C = 7 |
| **Mani A.R. et al. 2008 (13)** | To assess the relationship between indices of HRV, hepatic encephalopathy and systemic inflammation in cirrhotic patients | (1) Reduced long term HRV in cirrhotic patients compared with healthy subjects. (2) HRV negatively correlated with degree of neuropsychiatric impairment. (3) 8% increased relative risk of death for every 1ms drop in HRV index (4) Indices of HRV and neuropsychiatric performance significantly correlated with level of plasma IL6. | UK | inpatients (14)<br>outpatients (66) | 80(53) | 11 (5) | Alcohol = 65<br>HBV + HCV = 7<br>Mixed (Various) = 9 | A = 51 B = 13<br>C = 16 |
| **Milovanovic B. et al. 2009 (29)** | To analyse risk predictors of sudden cardiac death (SCD) related to autonomic dysfunction in alcohol-related cirrhotic patients. | (1) Patients with ARLD are susceptible to autonomic dysfunction (56%). (2) ARLD patients also have lower HRV indices (SDNN, SDANN, TINN, LF, and HF), serious arrhythmia, prolonged QTc and abnormal Poincare plot. (3) QTc inversely correlated with<br>lnLF, lnHF (r= -0.53, r= -0.47; p<0.05) while Lown class correlated with autonomic function (r= -0.64; p=0.05). | Serbia | inpatients | 25 (20) | 19 (15) | Alcohol = 25 | NS |
| **Miyajima H. et al. 2001 (30)** | To assess the effect of portal blood flow volume and autonomic nervous function on abnormal gastric motility in cirrhotic patients. | (1) Autonomic dysfunction correlated with abnormal gastric motility in cirrhotic patients (2) Decreased gastric motility may result from abnormalities in autonomic function in cirrhotic patients | Japan | NS | 27(19) | 20(13) | Alcohol = 1<br>HBV = 5<br>HCV = 21 | A = 7<br>B + C = 20 |
| **Moller S. et al. 2012 (31)** | To assess sympathetic control of cardiac function in cirrhosis using mIBG scintigraphy and relate this to cardiovascular functions | (1) Cirrhotic patients have significantly reduced HRV and baroreflex activity. (2) Reduction in HRV and baroreflex correlated significantly with abnormal cardiac sympathetic nervous activity measured by catecholamine uptake by mIBG. | Denmark | outpatients | 10 (5) | 10 (5) | Alcohol = 10 | NS |
| **Nagasako C.K. et al. 2009 (32)** | To assess autonomic dysfunction in nonalcoholic cirrhosis and the relationship with disturbed intestinal transit time, as well as severity and prognosis using HRV. | (1) HRV indices (HF, lnHF, LF, lnLF, pNN50) was lower in cirrhotic patients (Child B) patients compared with child A and control subjects. (2) HRV indices correlated significantly with the risk of hepatic encephalopathy in cirrhotic patients. | Brazil | NS | 32(12) | 21(12) | HCb = 16<br>PBC = 6<br>Cryptogenic = 6<br>Others = 4 | A = 13<br>B = 19<br>C = 0 |
| **Negru R.D. et al. 2015 (33)** | To assess the use of HRV as a marker of autonomic dysfunction and severity in cirrhotic patients. | (1) HRV detected autonomic dysfunction in cirrhotic patients. (2) Aetiology of liver cirrhosis is linked with different types of autonomic dysfunctions. (3) HRV parameters correlated significantly with severity of liver cirrhosis as assessed by ChildPugh scores. | Romania | inpatients | 52 (27) | 30 (15) | Alcohol = 25<br>HBV = 6<br>HCV = 17<br>Mixed = 4 | A = 30<br>B = 8<br>C = 14 |
| **Satti R. et al. 2019 (14)** | To extend the Poincare plot by introducing a sequential lag in the correlation computation and to evaluate the relationship with the severity and survival of cirrhotic patients. | (1) Traditional SD1 and SD2 correlated strongly with severity of liver cirrhosis. (2) Lagged SD1 and SD2 correlated significantly with liver disease severity. However, extended SD1 did not predict mortality independently of MELD. (3) SD2 predicted survival of cirrhotic patients independent of MELD. | Italy | Outpatients | 74 | 35 | NS | NS |

vs, versus; HRV, Heart Rate Variability; ARLD, Alcohol-Related Liver Disease; NALD, Non-Alcoholic Liver Disease (including virus-linked liver diseases); PBC, Primary Biliary Cholangitis; IL6, Interleukin-6; LT, Liver Transplantation; ln, Natural Logarithm; QTc, Q-T complex describing time Interval between the start of Q-wave and T-wave of an ECG recording; HCV, Hepatitis C Virus; MELD, Model for End-Stage Liver Disease; NN Interval, time lapse between consecutive QRS complexes of ECG recording; SDNN, Standard deviation of NN intervals; SDANN, Standard deviation of the average NN intervals for each 5-minute segments deduced from a 24-hour ECG recoding; pNN50, Percentage of successive RR intervals that vary by more than 50ms; RMSSD, Root mean square of differences in successive NN interval; TINN, Triangular Interpolation of the NN intervals' histogram; LF, Low Frequency; HF, High Frequency; SD1, Poincare plot Standard Deviation perpendicular to the line of identity; SD2, Poincare plot Standard Deviation along the line of identity; ApEn, Approximate Entropy; NS, Not Shown.

**Table 2.**
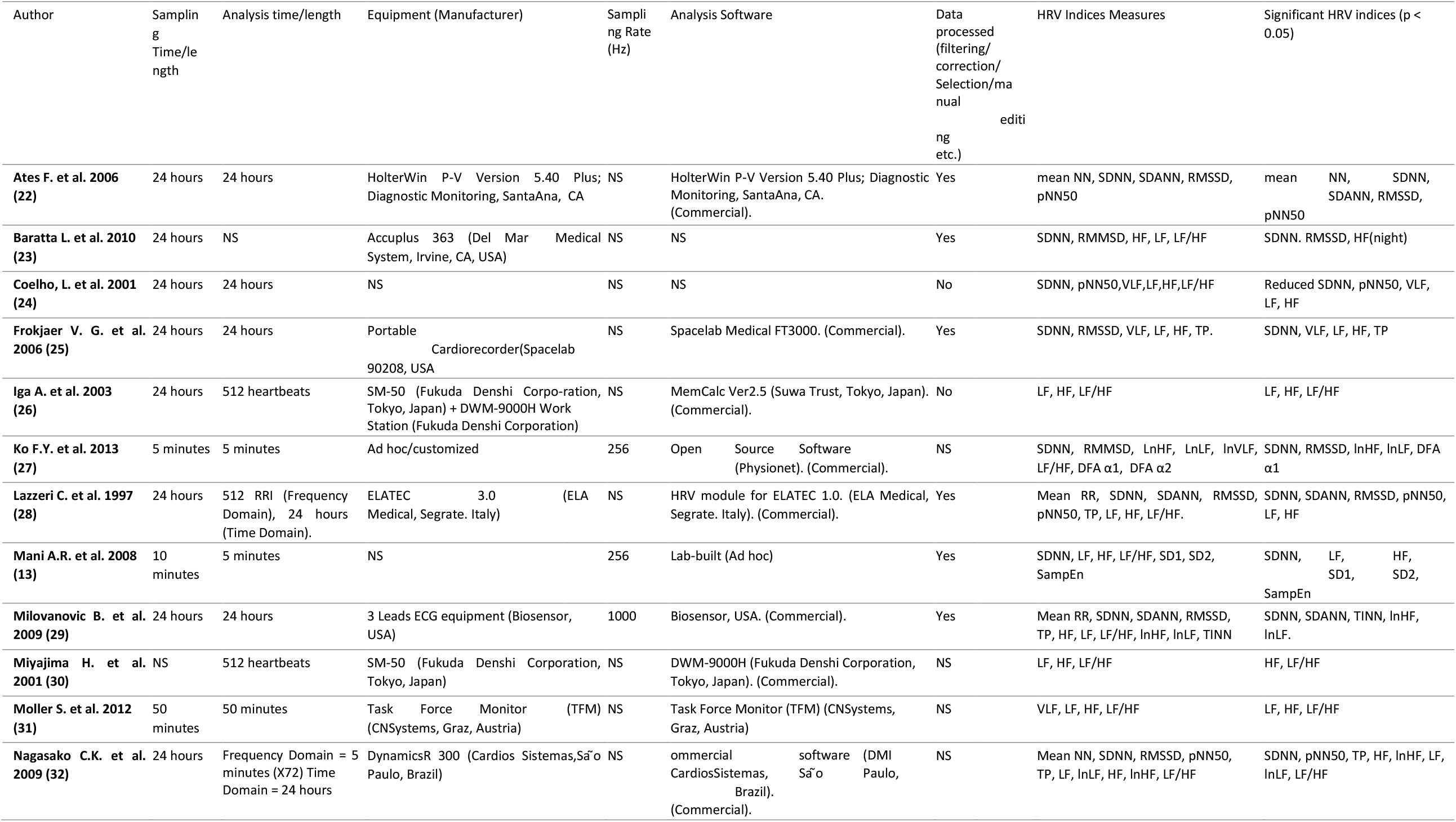
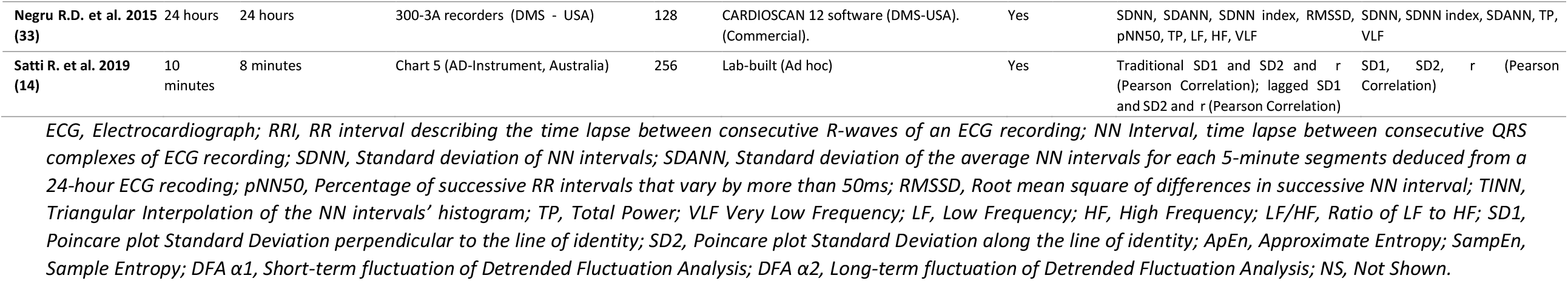
ECG recording and HRV analysis techniques of included studies.

For the survival prediction, studies that used HRV indices for survival analysis in cirrhosis were included in the mini-systematic review. Where reported, the sample size, follow-up time, mortality, HRV indices analysed, HRV indices that predicts mortality independently of MELD and Child-Pugh scores, HRV indices of survivors and non-survivors and hazard or odds ratios were extracted from the studies (Table 3).

**Table 3.**
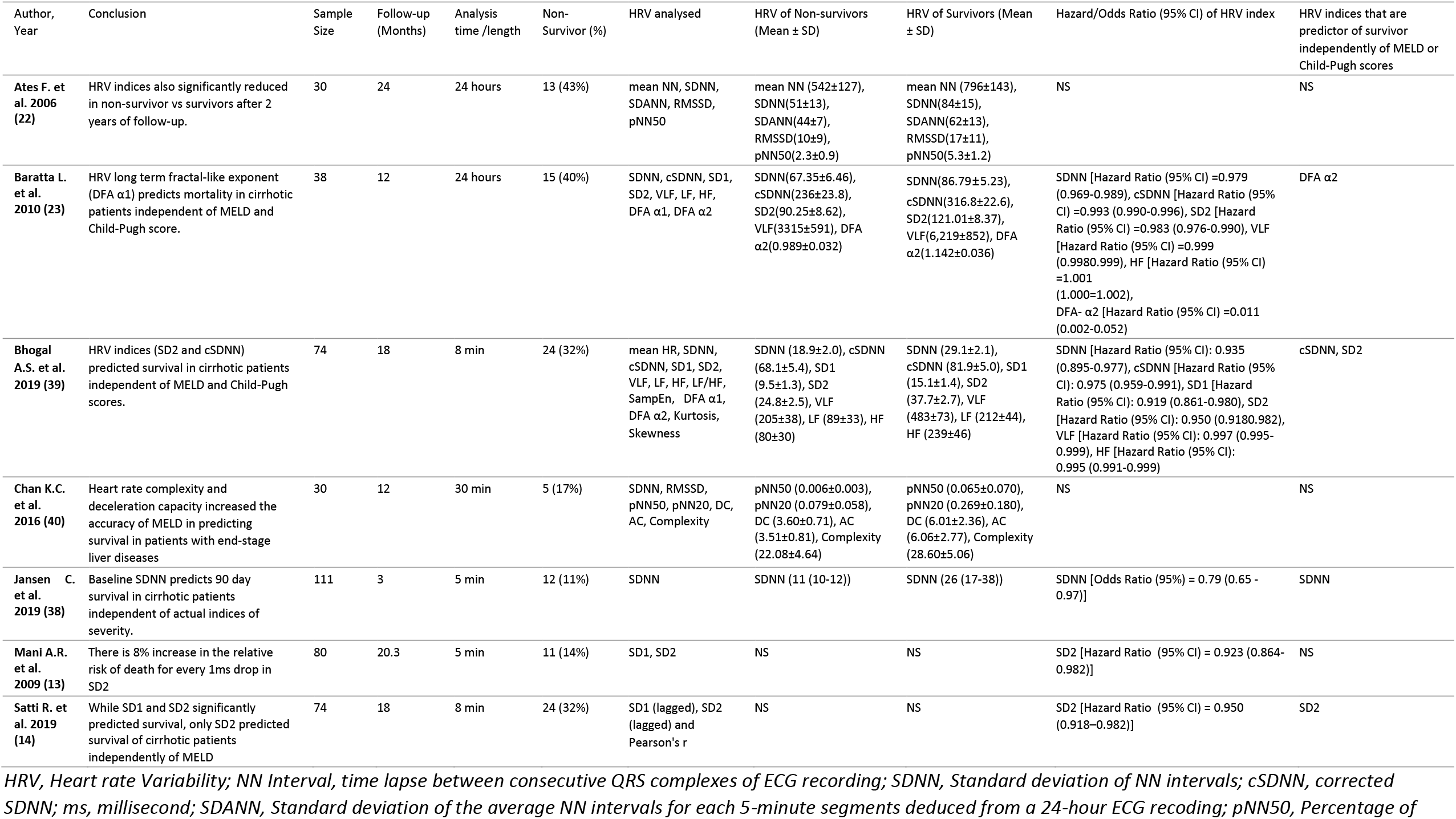
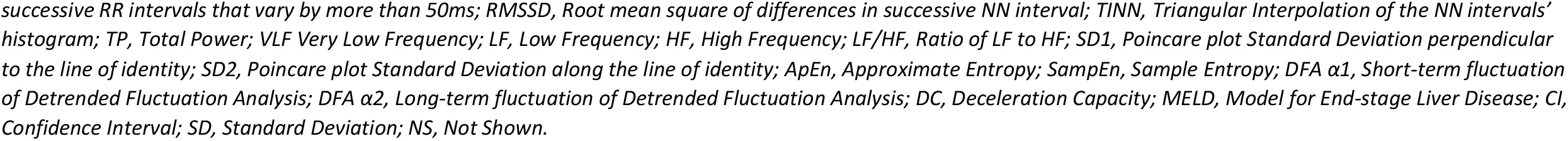
Characteristics of studies that reported the HRV indices as predictors of survival in patients with cirrhosis.

| Author, Year | Conclusion | Sample Size | Follow-up (Months) | Analysis time /length | Non-Survivor (%) | HRV analysed | HRV of Non-survivors (Mean ± SD) | HRV of Survivors (Mean ± SD) | Hazard/Odds Ratio (95% CI) of HRV index | HRV indices that are predictor of survivor independently of MELD or Child-Pugh scores |
| --- | --- | --- | --- | --- | --- | --- | --- | --- | --- | --- |
| **Ates F. et al. 2006 (22)** | HRV indices also significantly reduced in non-survivor vs survivors after 2 years of follow-up. | 30 | 24 | 24 hours | 13 (43%) | mean NN, SDNN, SDANN, RMSSD, pNN50 | mean NN (542±127), SDNN(51±13), SDANN(44±7), RMSSD(10±9), pNN50(2.3±0.9) | mean NN (796±143), SDNN(84±15), SDANN(62±13), RMSSD(17±11), pNN50(5.3±1.2) | NS | NS |
| **Baratta L. et al. 2010 (23)** | HRV long term fractal-like exponent (DFA α1) predicts mortality in cirrhotic patients independent of MELD and Child-Pugh score. | 38 | 12 | 24 hours | 15 (40%) | SDNN, cSDNN, SD1, SD2, VLF, LF, HF, DFA α1, DFA α2 | SDNN(67.35±6.46), cSDNN(236±23.8), SD2(90.25±8.62), VLF(3315±591), DFA α2(0.989±0.032) | SDNN(86.79±5.23), cSDNN(316.8±22.6), SD2(121.01±8.37), VLF(6,219±852), DFA α2(1.142±0.036) | SDNN [Hazard Ratio (95% CI) =0.979 (0.969-0.989), cSDNN [Hazard Ratio (95% CI) =0.993 (0.990-0.996), SD2 [Hazard Ratio (95% CI) =0.983 (0.976-0.990), VLF [Hazard Ratio (95% CI) =0.999 (0.9980-0.999), HF [Hazard Ratio (95% CI) =1.001 (1.000=1.002), DFA- α2 [Hazard Ratio (95% CI) =0.011 (0.002-0.052) | DFA α2 |
| **Bhogal A.S. et al. 2019 (39)** | HRV indices (SD2 and cSDNN) predicted survival in cirrhotic patients independent of MELD and Child-Pugh scores. | 74 | 18 | 8 min | 24 (32%) | mean HR, SDNN, cSDNN, SD1, SD2, VLF, LF, HF, LF/HF, SampEn, DFA α1, DFA α2, Kurtosis, Skewness | SDNN (18.9±2.0), cSDNN (68.1±5.4), SD1 (9.5±1.3), SD2 (24.8±2.5), VLF (205±38), LF (89±33), HF (80±30) | SDNN (29.1±2.1), cSDNN (81.9±5.0), SD1 (15.1±1.4), SD2 (37.7±2.7), VLF (483±73), LF (212±44), HF (239±46) | SDNN [Hazard Ratio (95% CI): 0.935 (0.895-0.977), cSDNN [Hazard Ratio (95% CI): 0.975 (0.959-0.991), SD1 [Hazard Ratio (95% CI): 0.919 (0.861-0.980), SD2 [Hazard Ratio (95% CI): 0.950 (0.9180-0.982), VLF [Hazard Ratio (95% CI): 0.997 (0.995-0.999), HF [Hazard Ratio (95% CI): 0.995 (0.991-0.999) | cSDNN, SD2 |
| **Chan K.C. et al. 2016 (40)** | Heart rate complexity and deceleration capacity increased the accuracy of MELD in predicting survival in patients with end-stage liver diseases | 30 | 12 | 30 min | 5 (17%) | SDNN, RMSSD, pNN50, pNN20, DC, AC, Complexity | pNN50 (0.006±0.003), pNN20 (0.079±0.058), DC (3.60±0.71), AC (3.51±0.81), Complexity (22.08±4.64) | pNN50 (0.065±0.070), pNN20 (0.269±0.180), DC (6.01±2.36), AC (6.06±2.77), Complexity (28.60±5.06) | NS | NS |
| **Jansen C. et al. 2019 (38)** | Baseline SDNN predicts 90 day survival in cirrhotic patients independent of actual indices of severity. | 111 | 3 | 5 min | 12 (11%) | SDNN | SDNN (11 (10-12)) | SDNN (26 (17-38)) | SDNN [Odds Ratio (95%) = 0.79 (0.65 - 0.97)] | SDNN |
| **Mani A.R. et al. 2009 (13)** | There is 8% increase in the relative risk of death for every 1ms drop in SD2 | 80 | 20.3 | 5 min | 11 (14%) | SD1, SD2 | NS | NS | SD2 [Hazard Ratio (95% CI) = 0.923 (0.864-0.982)] | NS |
| **Satti R. et al. 2019 (14)** | While SD1 and SD2 significantly predicted survival, only SD2 predicted survival of cirrhotic patients independently of MELD | 74 | 18 | 8 min | 24 (32%) | SD1 (lagged), SD2 (lagged) and Pearson's r | NS | NS | SD2 [Hazard Ratio (95% CI) = 0.950 (0.918–0.982)] | SD2 |

HRV, Heart rate Variability; NN Interval, time lapse between consecutive QRS complexes of ECG recording; SDNN, Standard deviation of NN intervals; cSDNN, corrected SDNN; ms, millisecond; SDANN, Standard deviation of the average NN intervals for each 5-minute segments deduced from a 24-hour ECG recoding; pNN50, Percentage of
*successive RR intervals that vary by more than 50ms; RMSSD, Root mean square of differences in successive NN interval; TINN, Triangular Interpolation of the NN intervals' histogram; TP, Total Power; VLF Very Low Frequency; LF, Low Frequency; HF, High Frequency; LF/HF, Ratio of LF to HF; SD1, Poincare plot Standard Deviation perpendicular to the line of identity; SD2, Poincare plot Standard Deviation along the line of identity; ApEn, Approximate Entropy; SampEn, Sample Entropy; DFA α1, Short-term fluctuation of Detrended Fluctuation Analysis; DFA α2, Long-term fluctuation of Detrended Fluctuation Analysis; DC, Deceleration Capacity; MELD, Model for End-stage Liver Disease; CI, Confidence Interval; SD, Standard Deviation; NS, Not Shown.*

### Quality assessment

Two authors independently assessed the quality of the methods used in the included studies. A modified version of the Newcastle Ottawa Scale (NOS) was used [33]. The assessment comprised 3 domains that evaluated selection, comparability and outcome techniques employed in the included studies. The domains have 6 subdomains (questions) each of which is scored with a star. Lower score (≤3 stars) were considered as having high risk of bias (Table S1).

### Data synthesis

A meta-analysis was computed to calculate the difference in HRV between patients with cirrhosis and healthy controls. Output was generated as forest plots of effect sizes of HRV indices between the groups using the *Metan* procedure in Stata/SE15. HRV indices reported as median and interquartile mode were transformed to mean and standard deviation according to (17, 18), while indices presented as natural log (LN) were transformed by finding exponents (e^x^) accordingly. For studies where day and night indices were reported, the data of the day were used.

The effect sizes in the form of standardized mean difference (SMD) of each of the reported indices were pooled using the Hedges’ criteria (19). Random-effect or fixed effect model were used depending on between-studies heterogeneity and effect sizes presented as SMD with 95% confidence intervals (CI) in HRV indices between the patients and heathy controls. The effect sizes were visualized as forest plots which include percentage weights and between-studies heterogeneity (I^2^ Statistic, p-value). The I^2^ statistic measures the degree of heterogeneity between pooled studies and can be putatively interpreted as low, moderate or high when values are 25%, 50% or 75% respectively (20). The SMDs were interpreted according to the recommendations of Hopkins et al. whereby values <0.2 = trivial; 0.2–0.6 = small, 0.6–1.2 = moderate, 1.2–2.0 = large, 2.0-4.0 = very large, and > 4.0 = extremely large effect sizes (21) (Figures 1 and 2).

**Figure 1.**
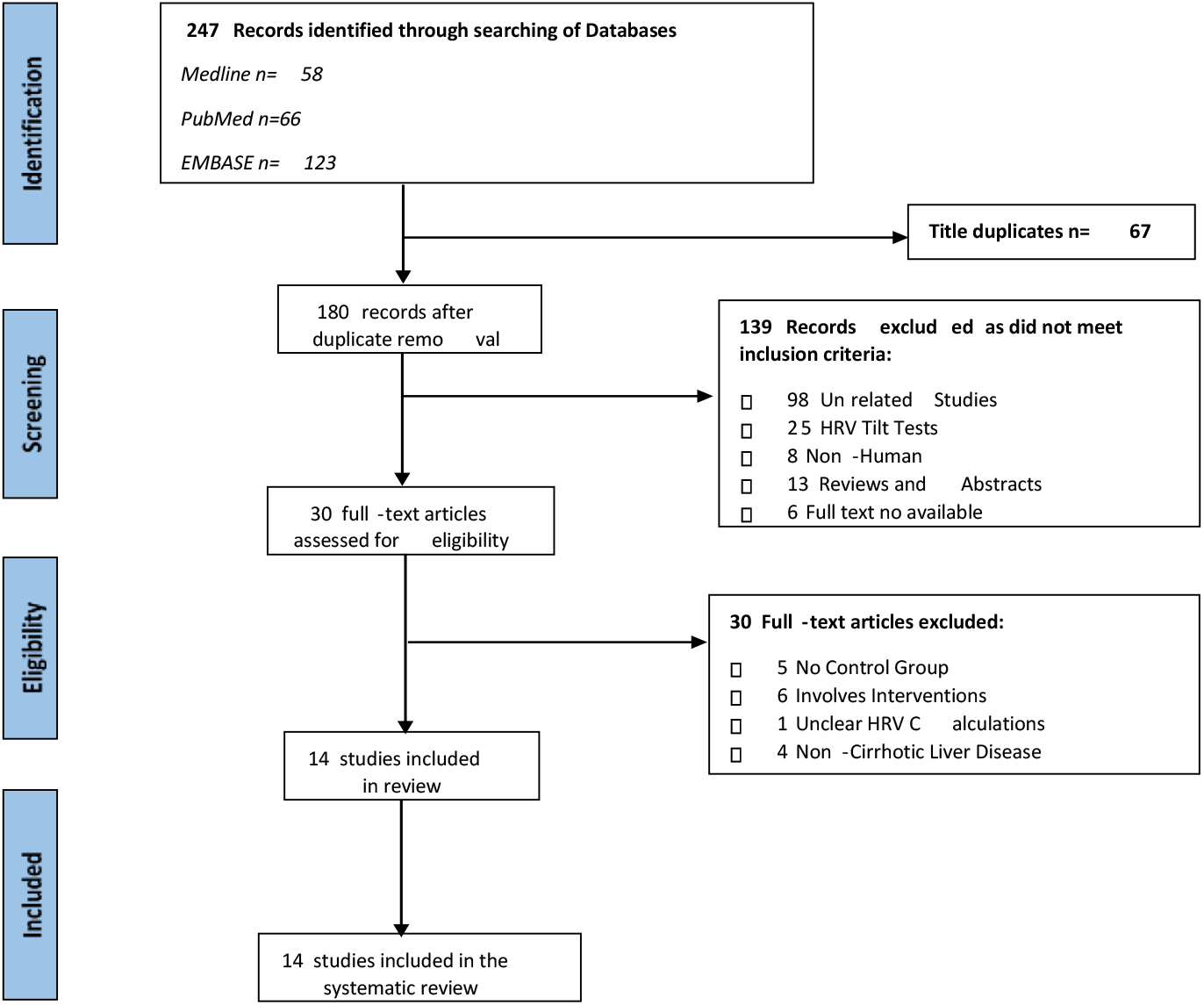
Heart rate variability in liver diseases, systematic review according to the Preferred Reporting Items for Systematic Reviews and Meta-analyses diagram.

**Figure 2(a, b, c and d).**
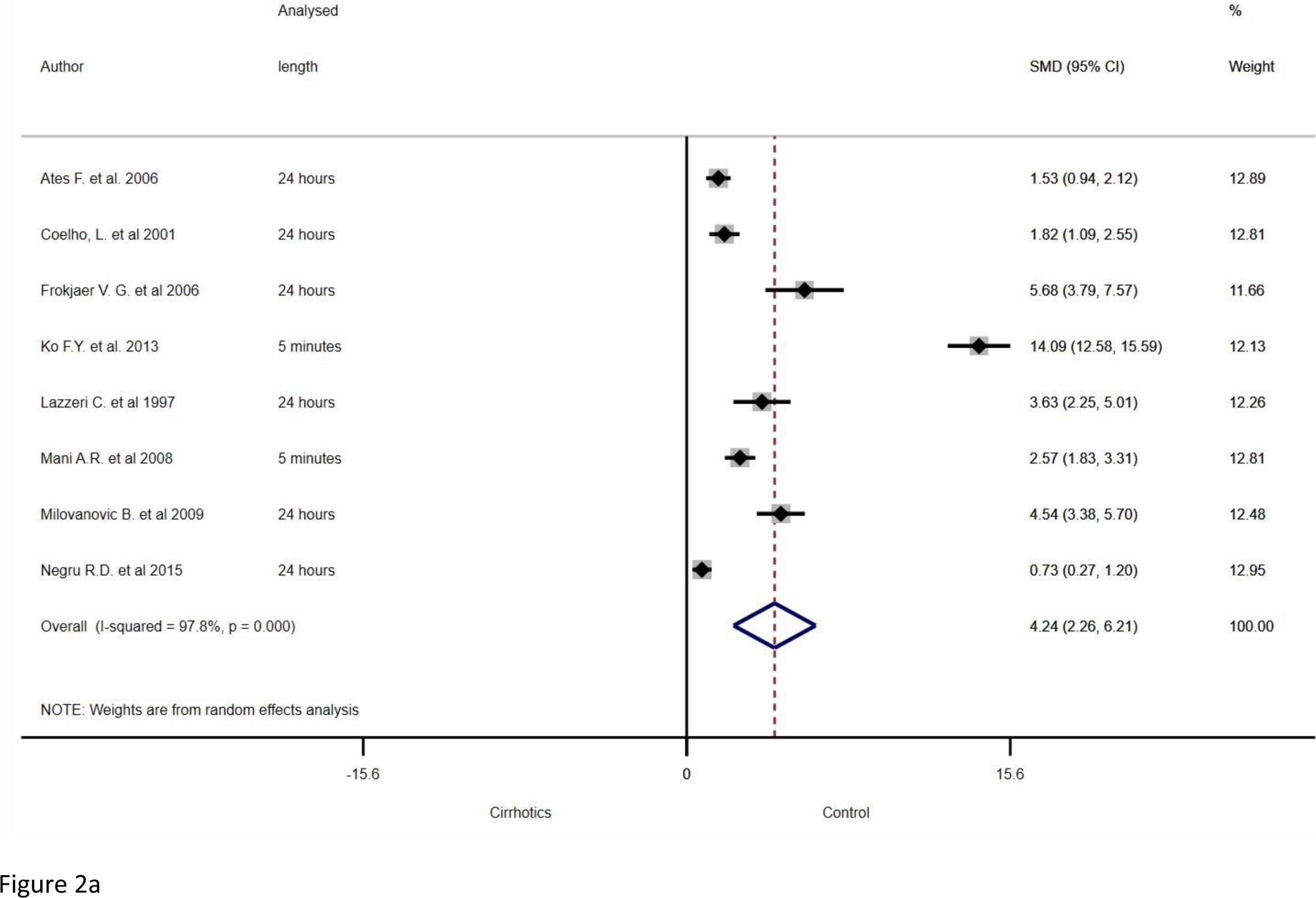
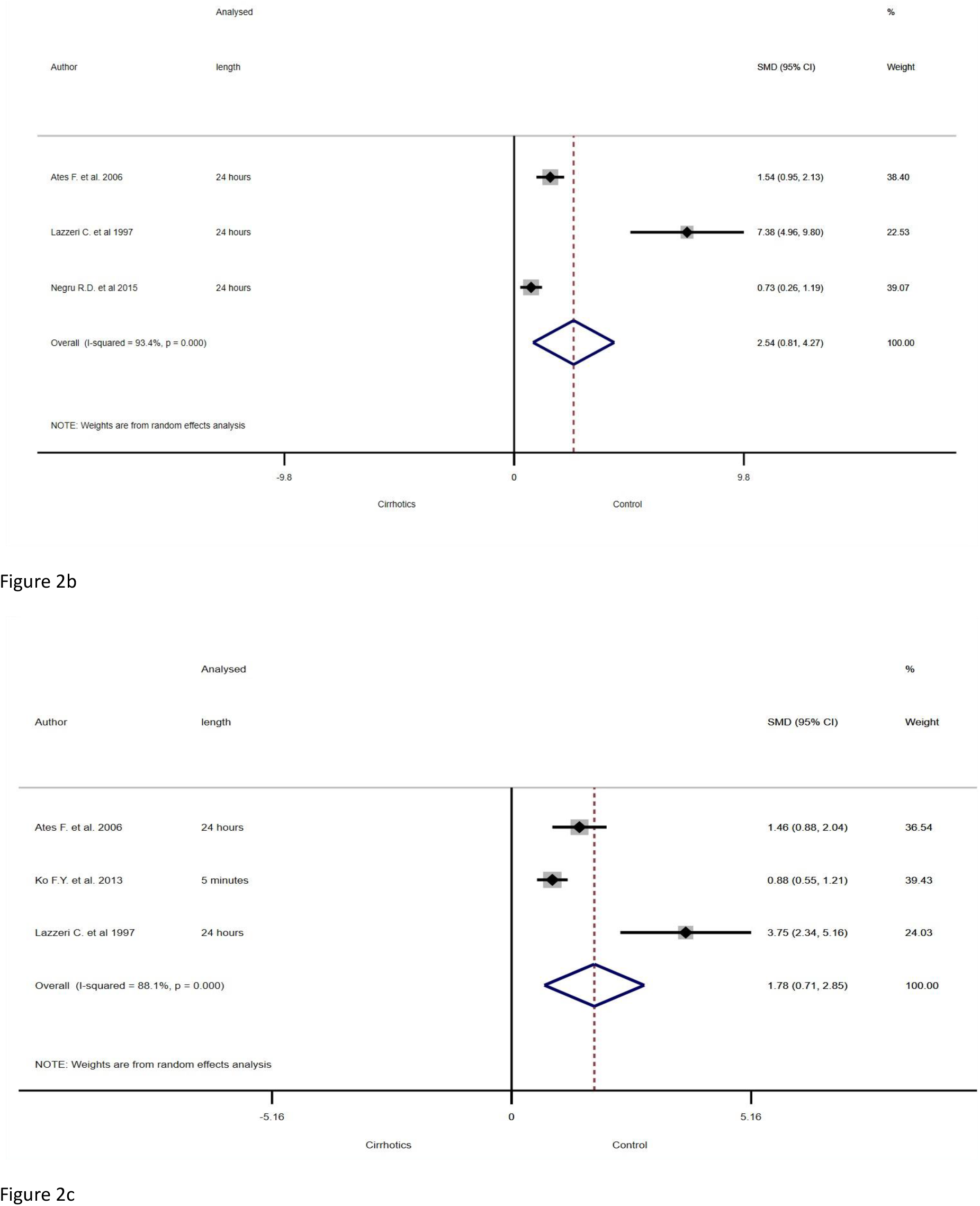
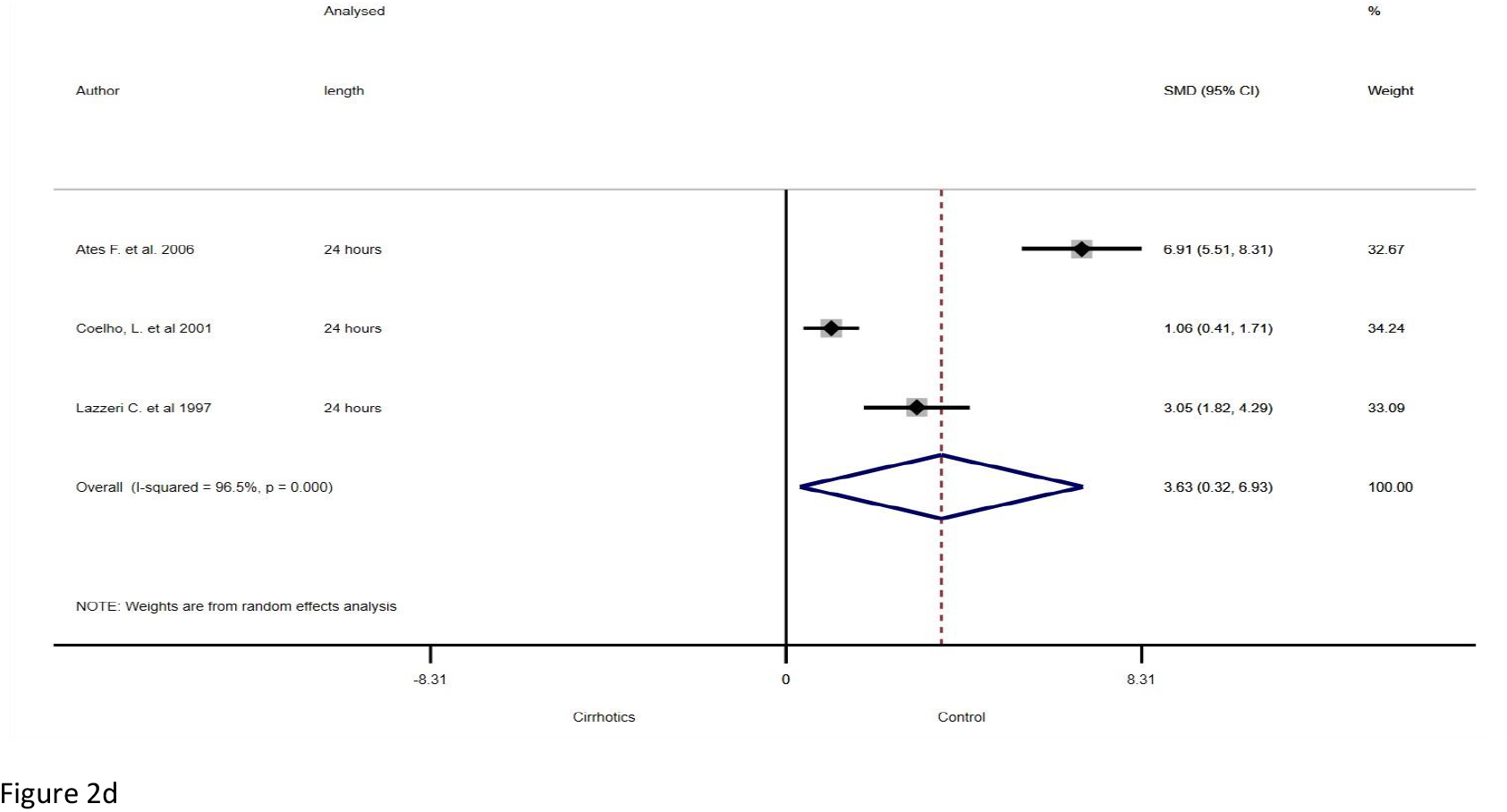
Forest plot for the standardized mean differences (SMD) in HRV time domain indices: SDNN (2a), SDANN (2b), RMSSD (2c) and pNN50 (2d) between patients with liver diseases and matched healthy controls. Hedges’ G effect size estimates were calculated with 95% confidence interval and computed using random effect model. Continuous horizontal lines and diamonds width represents 95% confidence interval and the diamonds centre and vertical red dotted line indicates the pooled random effect sizes.

Meta-analysis of the survival analysis was not performed because of the small number of studies found. Further, some of these studies originated from the same centre and possibly involved the same set of patients. Also, the indices reported varies between studies as well as the follow-up time.

## Results

Our database search generated a total of 247 studies of which 67 were duplicates. Of the remaining 180 studies, a total of 30 studies were deemed potentially relevant according to the inclusion criteria. However, a full text review resulted in the exclusion of a further 16 papers to give 14 studies that fulfilled all criteria (Figure 1). For the mini systematic review, a total of 7 studies compared the HRV indices of patients with cirrhosis that survived and did not survive.

### Description of included studies

Table 1 presents the general characteristics of included studies while Table 2 contains the techniques used for ECG recording, HRV analysis and indices reported to be significantly different between the groups. Table 4 presents the definitions for the HRV indices. The 14 included studies comprise a total of 583 patients and 349 healthy matched controls with sample size ranging between 20 and 180. All studies included are observational and conducted across 10 countries.

The risk of bias as assessed by the modified NOS scale showed that most studies have low risk of bias with none of the 14 studies scoring ≤ 3 (Table S1). Overall, most studies reported a reduction in HRV indices except LF-HF ratio (LF: HF) which was reported to be both increased [5, 10] and decreased [11] in cirrhosis. Table S2 contains definitions and units of the indices of HRV.

The mini-systematic review included 7 studies involving a total of 437 patients. Of these, 104 (24%) patients did not survive to the end of follow-up (3 to 24 months). All studies observed significant differences in HRV indices between the survivors and non-survivors (Table 3).

### HRV Time Domains

A total of 13 studies observed significant differences in HRV time domains between patients with cirrhosis and matched controls. The time domains reported include standard deviation of NN intervals (SDNN), SDNN Index, standard deviation of the average NN intervals for each 5 min segment of a 24 h ECG recording (SDANN), root mean square of successive NN interval (RMSSD) and percentage of NN intervals that differ by 50% (pNN50). Significant effect sizes were observed for each of the pooled time domain indices (SDNN, SDANN, RMSSD and pNN50%) which were significantly higher in the healthy controls. Overall, the betweenstudies heterogeneities were significantly high in all the pooled HRV time domains (Figures 2a-d).

#### SDNN

SDNN is the standard deviation of normal/non-ectopic RR intervals (NN) and is translated as the measure of the overall influence of autonomic nervous system on the variation of heart rhythm (34, 35). A total of 9 studies reported significant reduction in SDNN in cirrhosis (13, 22, 24, 25, 27-29, 32, 33). Eight of the 9 studies presented SDNN as mean (±SD) while one study (32) presented the median SDNN with no interquartile range (Table S3). This study was excluded from the effect size computation. A v*ery large* effect size was observed with significantly higher SDNN in healthy control compared with patients with cirrhosis [SMD (95%CI) = 3.41 (2.24, 4.58)]; Figure 2a]. This translates to noticeable dysregulation of the autonomic control of the cardiac rhythm in patients with cirrhosis.

#### SDNN Index

SDNN Index represents the average of all (288) 5 minutes-SDNNs of a 24-hour ECG recording. Physiologically, SDNN index has been linked with the overall autonomic control of heart rhythm (34, 35). Significant differences in the SDNN index was reported in only one of the included studies with a higher index and intact autonomic cardiac control in healthy controls (mean ± SD = 56.50 ± 17.04) compared with the patients (mean ± SD = 43.83 ± 15.66; Table S4) (33).

#### SDANN

SDANN is the standard deviation of all (288) averages of 5-minute NN intervals of a 24-hour ECG recording. SDANN is physiologically similar to SDNN in that it provides a measure of autonomic regulation of the heart rhythm (34, 35). Three studies reported a significant reduction in SDANN due to cirrhosis (Table S5) (22, 28, 33). A significantly higher SDANN was observed in healthy controls compared with patients with cirrhosis with a *very large* difference between the groups [SMD (95%CI) = 2.54 (0.81, 4.27); Figure 2b].

#### RMSSD

Root mean square of the NN intervals is the square root of the average of the square of NN intervals usually measured over 5 minutes. RMSSD has been linked with vagal influence on the heart rhythm and is used as an index of respiratory sinus arrhythmia (RSA) (34, 35). Overall, 3 studies reported significant alteration in RMSSD in the patients compared with healthy controls (Table S6) (22, 27, 28). There was significantly higher RMSSD in the control group with a *large* difference between the groups [SMD (95%CI) = 1.60 (0.73, 2.47); Figure 2c]. This can be interpreted as marked reduction in vagal control of the heart rhythm due to cirrhosis.

#### pNN50

Percentage of the NN intervals that differ from eachother by more than 50ms. The pNN50 indexes parasympathetic influence on heart rhythm and provides a comparatively less accurate assessment of RSA compared with the RMSSD (34, 35). Further, 4 of the pooled studies found significant reduction in pNN50 in cirrhosis compared with healthy control (22, 24, 28, 32). One of the studies reported median pNN50 without the interquartile range and was not included on the analysis (Table S7) (32). Indeed, lower RMSSD was reported in the patients compared with the control groups with a *very large* effect size between the group [SMD (95%CI) = 2.54 (1.21, 3.87); Figure 2d].

### HRV Frequency Domain

HRV Frequency domains represent the various frequency bands resulting from autoregressive (AR) or Fast Fourier Transformation (FFT) of the NN variations. Ten of the included studies reported significant difference in HRV frequency domains between patients with cirrhosis and healthy controls. Indices reported include total power (TP), high frequency (HF), low frequency (LF), very low frequency (VLF), and LF/HF ratio (HF: LF). The between-studies heterogeneity as measured by Chi-square was significantly high in pooled HF and LF (Figures 3b and 3c). Thus, random effect model was employed. Fixed effect model was used for pooling TP and VLF as the between studies heterogeneities were significantly low (Figures 3a and 3d).

**Figure 3(a, b, c, and d).**
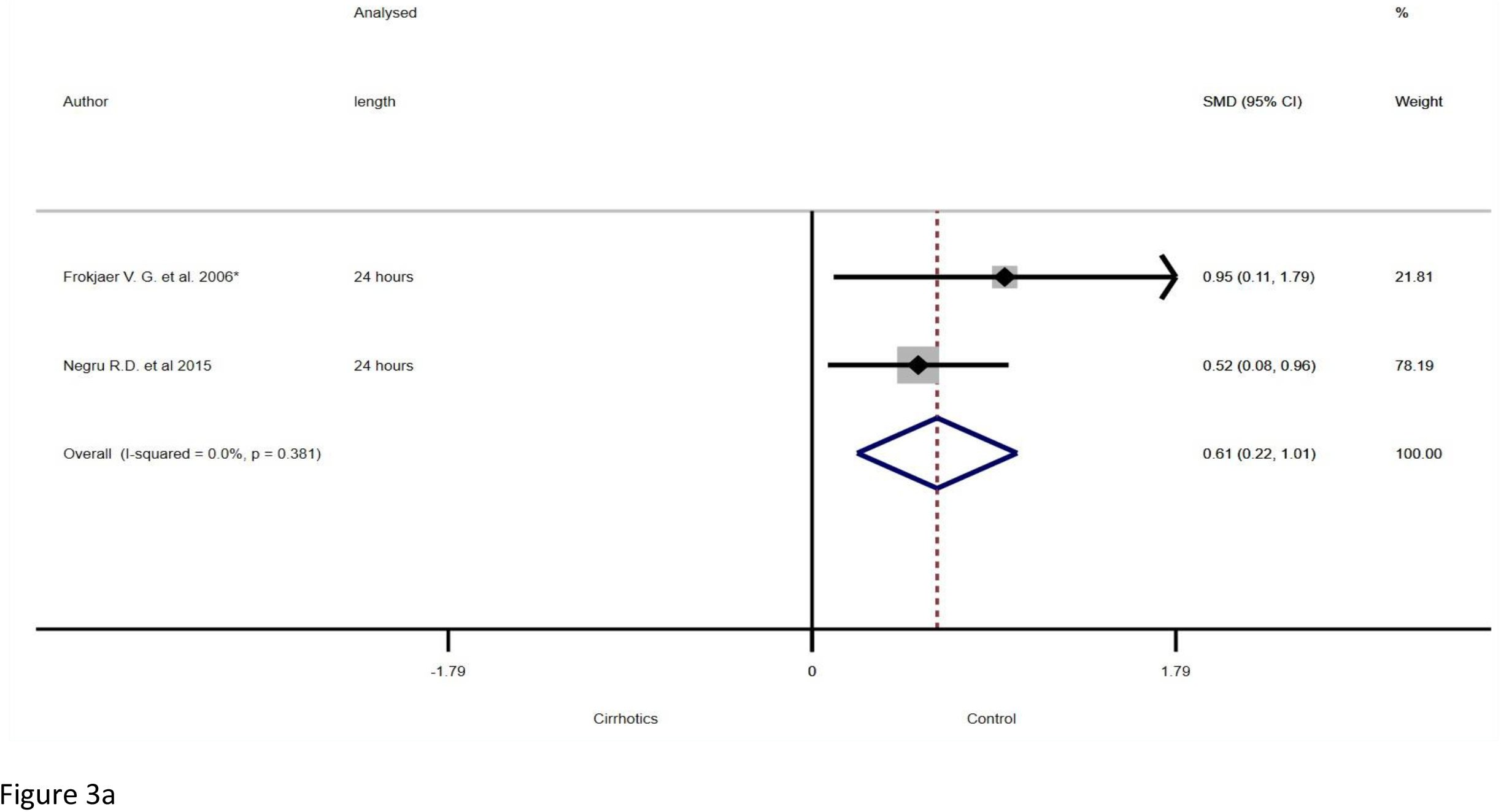
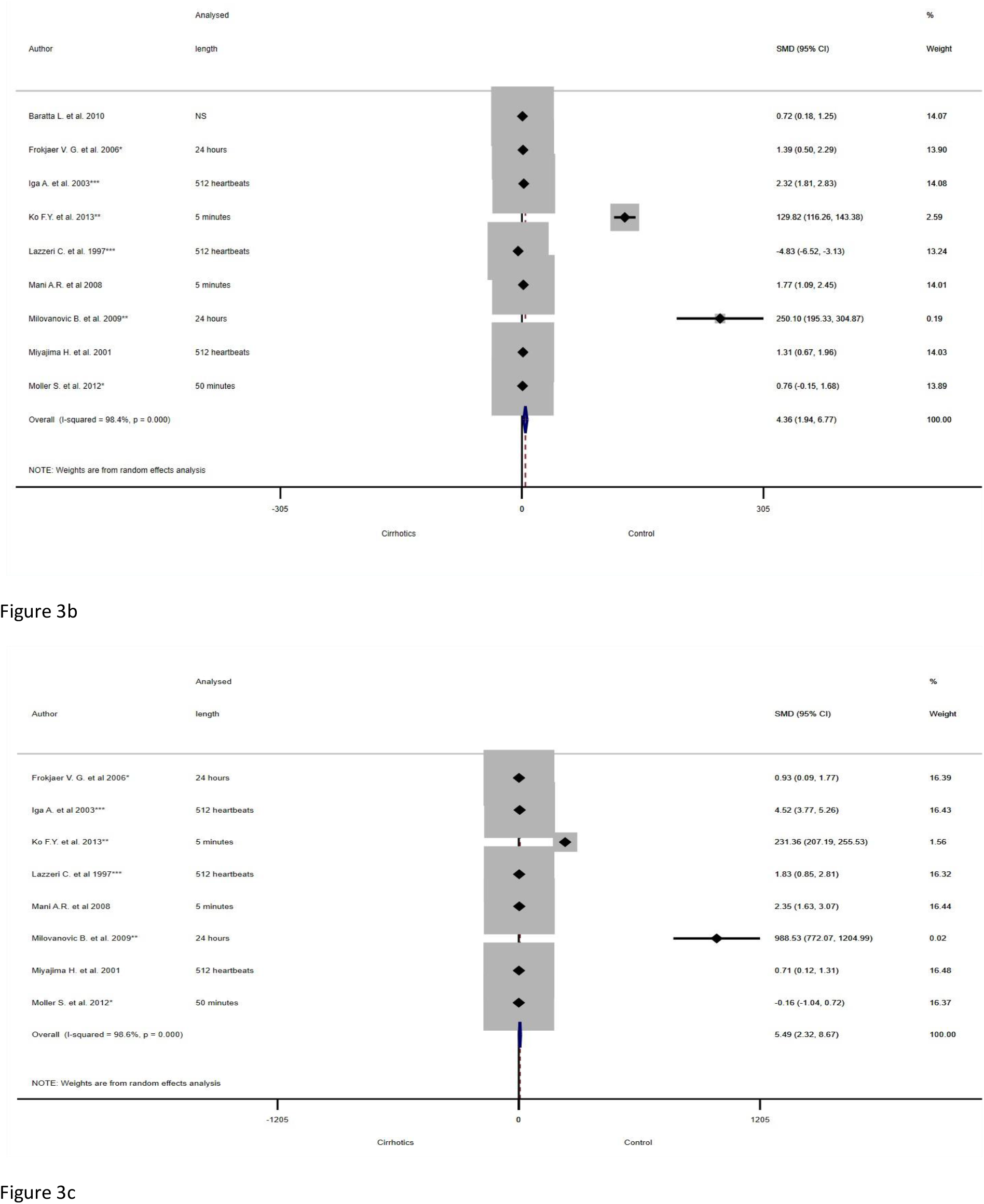
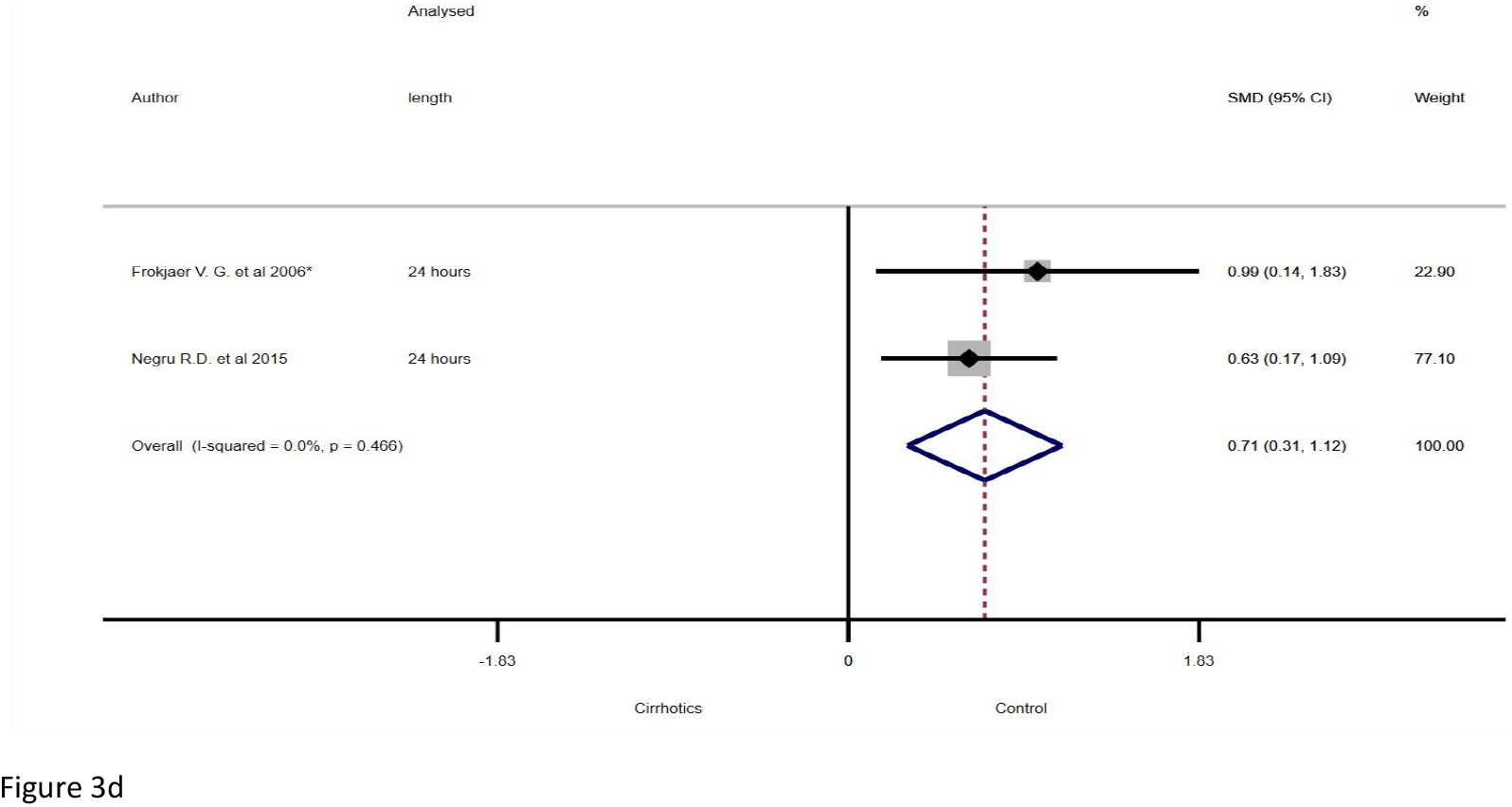
Forest plot for the standardized mean differences (SMD) in HRV frequency domain indices: TP (3a), HF (3b), LF (3c) and VLF (3d) between patients with liver diseases and matched healthy controls. Hedges’ G effect size estimates were calculated with 95% confidence interval and computed using random effect model. The width of the solid black diamonds represents 95% confidence interval of the effect sizes of each of the pooled studies and the blue diamond and vertical red dotted line indicates the pooled random or fixed effect sizes.

#### Total Power (TP)

Total power represents the aggregate energy in all the frequency bands (ULF, VLF, LF, and HF) of an ECG recording (34, 35). Two studies reported significantly reduced TP in cirrhosis (25, 33). All reported TP were analysed for 24 hours ECG recordings and reported as natural log in one of the included studies (25) (Table S8). There was an overall higher TP observed in controls compared with the patients. A *moderate* effect size was observed between the group [SMD (95%CI) = 0.62 (0.23, 1.02); Figure 3a].

#### High Frequency (HF)

High frequency is the power within the 0.15Hz – 0.4Hz frequency band of an ECG recording. The HF represents the parasympathetic control of the heart rhythm and the variation linked with the respiratory sinus arrhythmia (RSA) (34, 35). A total of 9 studies reported significant difference in HRV high frequency between the patients and controls (13, 23, 25-31). Of these, 8 studies reported reduced HF while 1 study reported an increased HF in cirrhosis (28) (Table S9). This study was not included in the data analysis because the model used does not correct for the difference in direction of effects (36). HF in healthy controls was observed to be higher compared with the patients, with an “extremely large” effect size between the groups [SMD (95%CI) = 4.36 (1.94, 6.77); Figure 3b]. This translate as an easily discernible dysregulation in vagal control of the heart rhythm in cirrhosis.

#### Low Frequency (LF)

Low frequency domain represents the power within 0.04–0.15 Hz frequency band of an ECG recording. The LF correlates strongly with baroreflex influence on the heart rhythm and is associated with both sympathetic and parasympathetic nervous controls (34, 35). Eight studies reported significantly different LF between the patients and healthy controls (Table S10) (13, 25-29, 31). Low frequency power was observed to be significanlty lower in cirrhosis compared with control with an *extremely large* difference observed in the effect size [SMD (95%CI) = 5.49 (2.32, 8.67); Figure 3c]. Thus, a marked reduction in response of the heart rhythm to the the baroreflex loop which may be attributed to cirrhosis.

#### Very Low frequency (VLF)

Very low frequency represents the power in the 0.0033–0.04 Hz frequency band of an ECG recording. While uncertainty exists in the physiological factors responsible for the VLF band, it has been strongly linked with the renin-angiotensin system, thermoregulation and endothelial factors (34, 35). Two studies reported significant difference in 24-hour VLF between healthy controls and cirrhotic patients (Table S11) (25, 33). Healthy control group were observed to have significantly higher VLF compared with patients with cirrhosis. The effect size for VLF between the group was moderate [SMD (95%CI) = 0.73 (0.32, 1.13); Figure 3d]. This can be translated as a reasonably observable stronger response of the heart rhythm to renin-angiotensin system, thermoregulation and endothelial factors in healthy controls compared with the patients with cirrhosis.

#### Low Frequency – High Frequency Ratio (LF: HF)

The ratio of low frequency power to high frequency power of an ECG recording. The LF: HF is traditionally translated as the measure of the sympatho-vagal balance because LF and HF had been hypothesised as measures of purely sympathetic and parasympathetic cardiac controls respectively (34, 35). However, this notion was challenged when it was observed that LF is influenced by both arms of the autonomic nervous system (37). A total of 3 studies reported significant difference in LF: HF between the patients and controls (26, 30, 31). Of these, 2 studies reported increase LF:HF in patients with cirrhosis (26, 30) and were pooled (Table S12). However, no significant effect size exist between the groups [i.e. test that SMD = 0: z = 2.89 p = 0.004].

### HRV Non-linear Indices

A total of 2 studies reported significant difference in non-linear HRV indices between patients with cirrhosis and healthy controls (13, 27). Four non-linear indices were reported including short-term (SD1) and longterm (SD2) HRV extracted from the Poincare’ plot, Sample Entropy and scaling exponent (α) calculated using detrended fluctuation analysis (DFA). Short-term and long-term variability (SD1 and SD2) of Poincare plot as well as Sample Entropy was reported to be significantly reduced in cirrhosis in one study (Table S13) (13). The short-term scaling exponent (DFA α1) which indicates the fractal-like pattern of cardiac rhythm was also reported to be altered in cirrhosis compared with healthy controls in one study [6].

### HRV in Survival Analysis

An overall of 7 studies assessed whether HRV can predicts survival in patients with cirrhosis for a follow-up period between 3 to 24 months (13-15, 22, 38-40). All studies concluded that HRV indices were significantly different between survivors and non-survivors. Indeed, based on the hazard or odds ratios reported, increased DFA α2, SD2, cSDNN, SDNN and VLF were significantly correlated with increased survival (Table 3). Of the 7 studies, 4 reported that HRV indices including DFA α2, cSDNN (corrected SDNN), SD2, and SDNN may predict mortality in cirrhosis independent of MELD and/or Child-Pugh scores.

## Discussion

We report here the effect of cirrhosis on autonomic cardiac regulation measured by indices of heart rate variability. Significant difference in HRV was observed between patients with cirrhosis and healthy controls, albeit between-studies heterogeneities were high. To address the heterogeneity of included studies, we used mostly random-effect model and pooled the standardized mean difference (SMD). In most cases, the HRV indices indicated autonomic dysfunction as shown previously (41). Some HRV indices (DFA α2, SD2, cSDNN and SDNN) also exhibited a significant correlation with severity of cirrhosis and survival of patients. Precisely, there was significant reduction in HRV time and frequency domain indices including SDNN, SDNN index, SDANN, RMSSD, pNN50 as well as TP, HF, LF and VLF in cirrhosis which correlated with disease severity. The relationship between cirrhosis and the ratio of LF: HF, which traditionally represent sympathovagal cardiac regulation is not clear with one study reporting increased (31) and two studies reporting decrease in cirrhosis (26, 30).

Importantly, the mini-systematic review shows that some indices of HRV predict survival in cirrhosis independently of measures of severity (MELD score). This is consistent with a recent report whereby SDNN was shown to independently predict mortality in patients with decompensated cirrhosis. Further, Jansen et al. also showed that SDNN was significantly reduced and correlated with increased plasma level of inflammatory biomarkers and severity of cirrhosis (38). Indeed, the association between cardiac autonomic dysregulation and systemic inflammation has been reported in cirrhosis (13) as well as in other diseases (42, 43). Similarly, the development of acute on chronic liver failure (ACLF) has been extensively linked with systemic inflammation (44-46). Thus, the suggestion that sudden reduction in HRV can be used to detect dynamic changes indicative of early acute decompensation in patients with cirrhosis (38). However, knowledge on the mechanism of cardiac autonomic dysfunction during systemic inflammation have only be reported in animal models and awaits further investigation in humans (47-50). Also, data are emerging to show that HRV correlates with sub-clinical hepatic encephalopathy (13), and may therefore become an indirect means to identify patients most likley to have sub-clinical hepatic encephalopathy without the need for an EEG (51). Perhaps, HRV may provide a relatively simpler and portable yet effective assessment of covert hepatic encephalopathy.

Reduction in HRV is not unique to cirrhosis and have been reported in non-cirrhotic liver patients. For example, Keresztes et al. reported significant reduction in HRV time and frequency domains in patients with primary biliary cholangitis compared with age-matched healthy controls. Indeed, 58% of the PBC patients studied also had autonomic dysfunction with abnormal cardiovascular reflex tests (52). Further, patients with chronic hepatitis C infection have impaired autonomic function assessed by HRV time and frequency indices, and this correlates with the degree of liver injury as assessed by serum alanine aminotransferase levels (53).

This study also observed that non-linear HRV indices were significantly impaired in cirrhosis. Short and long term HRV, measured by SD1 and SD2 respectively and sample entropy were reduced in cirrhosis compared with healthy controls. Our findings substantiate the report by Bhogal et al. in which SD2 as well as cSDNN were found to predict mortality independent of MELD in patients with cirrhosis (39). The long-term fractallike scaling exponent (DFA α2) was also lower in patients with cirrhosis predicts mortality independent of the measures of severity of liver failure (13). Non-linear HRV indices are measures of unpredictability and provides an index of complexity in inter-beats intervals. Increased complexity may be interpreted physiologically as increased flexibility and inclination of the cardiac rhythm to respond to environmental changes and autonomic nervous control. Indeed, increased memory length of cardiac rhythm, which can be interpreted as reduced physiological controllability, has been reported in patients with liver cirrhosis compared to healthy controls (54). Likewise, we recently showed that heart rate turbulence onset (TO) following premature ventricular contraction, a phenomenon that is linked with autonomic nervous control is reduced and predicts survival in cirrhosis (55). Put together, while the mechanistic link remain unclear, autonomic regulation of the cardiac rhythm is dysregulated in cirrhosis and may improve clinical diagnosis and prognosis of patients. Finally, while lower HRV complexity (Sample Entropy) have been reported in cirrhosis, its association with poor outcome has not been reported and should be a focus of future investigation.

This study has several limitations. Because of the difference in the time of ECG recording in the studies, it was not possible to assess the role of circadian rhythm in HRV. Further, few studies reported non-linear indices of HRV, and thus data could not be pooled to assess the differences or the survival analysis because of the variability in the follow-up time.

We conclude that HRV analysis has the potential to be applied in both clinical and research settings for the evaluation of autonomic dysfunction in patients with cirrhosis, and may be of value in the early detection of subclinical deterioration such as early covert hepatic encephalopathy or acute on chronic liver decompensation. Further, HRV is an independent predictor of outcome of cirrhosis and may improve the predictive power of MELD and Child-Pugh. Finally, despite the potential, variation in techniques of HRV measurement remain the main barrier to useful interpretation and applicability. Indeed, to generate robust data, there needs to be standardisation of techniques including ECG recording and HRV measurement in future. We hope that this paper provides an impetus to agreed standardisation of methodology to enable this field to develop.

## Data Availability

All relevant data related to this study is included or uploaded as supplemental material.

