## Supplemental Tables and Figures for "Heart Rate Variability in Patients with Cirrhosis: A Systematic Review and Meta-analysis"

**Supplementary Data**

**Table S1.** Risk of bias assessment of included studies using modified Newcastle-Ottawa scale

| **Studies** | **Selection** | | **Comparability** | | | **Outcome** | **Total**  (≤3 Stars = High Risk of Bias) |
| --- | --- | --- | --- | --- | --- | --- | --- |
|  | Random recruitment of patients and healthy control | Diagnosis of CLD | Age-matched | Sex-matched | Patient with diseases, treatment or lifestyle that influences HRV excluded | MELD or Child-Pugh reported |  |
| Ates F. et al. 2006 [1] | * |  | * | * | * | * | ***** |
| Baratta L. et al. 2010 [2] | * |  | * | * | * | * | ***** |
| Coelho, L. et al. 2001 [3] | * |  | * | * |  | * | **** |
| Frokjaer V. G. et al. 2006 [4] | * | * | * |  | * | * | ***** |
| Iga A. et al. 2003 [5] | * | * | * | * | * | * | ****** |
| Ko F.Y. et al. 2013 [6] | * | * | * |  | * | * | ***** |
| Lazzeri C. et al. 1997 [7] | * | * | * | * | * | * | ****** |
| Mani A.R. et al. 2008 [8] | * | * | * |  | * | * | ***** |
| Milova vic B. et al. 2009 [9] | * | * | * | * | * |  | ***** |
| Miyajima H. et al. 2001 [10] | * | * | * | * |  | * | ***** |
| Moller S. et al. 2012 [11] | * | * | * | * | * | * | ****** |
| Nagasako C.K. et al. 2009 [12] | * | * | * |  | * | * | ***** |
| Negru R.D. et al. 2015 [13] | * | * | * | * | * | * | ****** |
| Satti R. et al. 2019 [14] | * | * | * |  |  | * | **** |

*CLD, Chronic Liver Disease; HRV, Heart Rate Variability; MELD, Model for End-stage Liver Disease*

**Table S2.** Definitions and units of included HRV indices

| **HRV Indices** | **Units** | **Descriptions** |
| --- | --- | --- |
| NN intervals. | ms | Time difference between consecutive normal QRS complexes of an ECG recording. |
| SDNN | ms | Standard deviation of NN intervals. |
| cSDNN | ms | Corrected SDNN is the standard deviation of NN interval that have been corrected for heart rate. It is calculated as SDNN/e^–(HR/58.8)^ [see (15)]. |
| SDANN | ms | Standard deviation of the average NN intervals for each 5-minute segments deduced from a 24-hour ECG recording. |
| SDNN Index | ms | Mean of standard deviations of all NN intervals for each 5-minute segments deduced from a 24-hour ECG recording. |
| pNN50 | % | Percentage of successive NN intervals that vary by more than 50ms. |
| RMSSD | ms | Root mean square of differences in successive NN interval. |
| TINN | ms | Width of the base of a computed RR interval histogram. |
| TP |  | Variance of entire NN interval of either 5 minutes (short) or 24-hour (long) ECG recording. |
| VLF | ms^2^ | Power of the frequency band between 0.0033-0.04 Hz. |
| LF | ms^2^ | Power of the frequency band between 0.04-0.15 Hz. |
| HF | ms^2^ | Power of the frequency band between 0.15-0.4 Hz. |
| LF/HF |  | Ratio of LF power to HF power. |
| SD1 | ms | The length of the line or standard deviation perpendicular to the line of identity of a Poincare plot |
| SD2 | ms | The length of the line or standard deviation parallel to the line of identity of a Poincare plot |
| ApEn (Approximate Entropy) |  | Measure of irregularity and complexity of a series of NN intervals. |
| SampEn (Sample Entropy) |  | Measure of irregularity and complexity of a series of NN intervals. |
| DFA α1 (Short-term scaling exponent) |  | Measure of short-term fractal-like fluctuations of inter-beat intervals. |
| DFA α2 (Long-term scaling exponent) |  | Measure of long-term fractal-like fluctuations of inter-beat intervals. |

*ECG, Electrocardiograph; NN Interval, time lapse between consecutive QRS complexes of ECG recording; SDNN, Standard deviation of NN intervals; ms, millisecond; SDANN, Standard deviation of the average NN intervals for each 5-minute segments deduced from a 24-hour ECG recoding; pNN50, Percentage of successive RR intervals that vary by more than 50ms; RMSSD, Root mean square of differences in successive NN interval; TINN, Triangular Interpolation of the NN intervals’ histogram; TP, Total Power; VLF Very Low Frequency; LF, Low Frequency; HF, High Frequency; LF/HF, Ratio of LF to HF; SD1, Poincare plot Standard Deviation perpendicular to the line of identity; SD2, Poincare plot Standard Deviation along the line of identity; ApEn, Approximate Entropy; SampEn, Sample Entropy; DFA α1, Short-term fluctuation of Detrended Fluctuation Analysis; DFA α2, Long-term fluctuation of Detrended Fluctuation Analysis; ms, millisecond.*

**Table S3 –** Reported SDNN of included studies.

| **Author** | **Normal SDNN (Mean±SD) (ms)** | **Abnormal SDNN (Mean±SD) (ms)** |
| --- | --- | --- |
| Ates F. et al. 2006 [1] | 134 ± 52 | 70 ± 28 |
| Coelho, L. et al. 2001 [3] | 148.9 ± 33.97 | 84.14 ± 35.78 |
| Frokjaer V. G. et al. 2006 [4] | 139 ± 13 | 72 ± 10 |
| Ko F.Y. et al. 2013 [6] | 42.9 ± 15.1 | 22.8 ± 13.8 |
| Lazzeri C. et al 1997 [7] | 57.99 ± 7 | 36.55 ± 4 |
| Mani A.R. et al. 2008 [8] | 36.9 ± 14.2 | 14.4 ± 7.7 |
| Milovanovic B. et al 2009 [9] | 463.15 ± 111.83 | 93.42 ± 42.69 |
| Nagasako C.K. et al. 2009*** [12] | 79 | 56 |
| Negru R.D. et al. 2015 [13] | 129.60 ± 45.70 | 101.88 ± 31.79 |

***=Reported as natural log; ***=Reported as median, no interquartile range. All other indices are reported as mean ± standard deviation.*

*SDNN; Standard deviation of NN intervals; SD, Standard Deviation; ms, Millisecond.*

**Table S4 –** Reported SDNN Index of included studies

| **Author** | **Normal SDNN index (Mean±SD) (ms)** | **Abnormal SDNN index (Mean±SD) (ms)** |
| --- | --- | --- |
| Negru R.D. et al. 2015 [13] | 56.50 ± 17.04 | 43.83 ± 15.66 |

*SDNN; Standard deviation of NN intervals; SD, Standard Deviation; ms, Millisecond.*

**Table S5 –** Reported SDANN of included studies.

| **Author** | **Normal SDANN (Mean±SD) (ms)** | **Abnormal SDANN (Mean±SD) (ms)** |
| --- | --- | --- |
| Ates F. et al. 2006 [1] | 118 ± 49 | 55 ± 30 |
| Lazzeri C. et al. 1997 [7] | 125.57 ± 8 | 68.14 + 7 |
| Negru R.D. et al. 2015 [13] | 114.80 ± 43.48 | 88.21 ± 31.48 |

*All indices are reported as mean ± standard deviation.*

*SDNN; SDANN, Standard deviation of the average NN intervals for each 5-minute segments deduced from a 24-hour ECG recoding; SD, Standard Deviation; ms, Millisecond.*

**Table S6 –** Reported RMSSD of included studies.

| **Author** | **Normal RMSSD (Mean±SD) (ms)** | **Abnormal RMSSD (Mean±SD) (ms)** |
| --- | --- | --- |
| Ates F. et al. 2006 [1] | 39±22 | 14±10 |
| Ko F.Y. et al. 2013 [6] | 28.2 ± 10.1 | 18.3 ± 11.7 |
| Lazzeri C. et al. 1997 [7] | 41 ± l | 27 ± 5 |

***= Reported as natural log. All other indices are reported as mean ± standard deviation.*

*RMSSD, Root mean square of differences in successive NN interval; SD, Standard Deviation; ms, Millisecond.*

**Table S7 –** Reported pNN50 of included studies.

| **Author** | **Normal pNN50 (Mean±SD) (%)** | **Abnormal pNN50 (Mean±SD) (%)** |
| --- | --- | --- |
| Ates F. et al. 2006 [1] | 8.2 ± 0.6 | 4.0 ± 0.6 |
| Coelho, L. et al. 2001 [3] | 11.17 ± 9.88 | 3.54 ± 4.61 |
| Lazzeri C. et al. 1997 [7] | 13 ± 2 | 3 ± 4 |
| Nagasako C.K. et al. 2009*** [12] | 15.5 | 3.3 |

****= Reported as median, no interquartile range. All other indices are reported as mean ± standard deviation.*

*pNN50, Percentage of successive RR intervals that vary by more than 50 ms; SD, Standard Deviation; ms, Millisecond.*

**Table S8 –** Reported Total Power (TP) of included studies.

| **Author** | **Normal TP (Mean±SD) (ms^2^)** | **Abnormal TP(Mean±SD) (ms^2^)** |
| --- | --- | --- |
| Frokjaer V. G. et al 2006** [4] | 2307 ± 3258.1 | 204 ± 144.2 |
| Negru R.D. et al. 2015 [13] | 2724.74 ± 1530.04 | 2007.70 ± 1247.49 |

***= Reported as natural log. All other indices are reported as mean ± standard deviation.*

*TP, Total Power; SD, Standard Deviation; ms, Millisecond.*

**Table S9 –** Reported High Frequency (HF) of included studies.

| **Author** | **Normal HF (Mean±SD) (ms^2^)** | **Abnormal HF (Mean±SD) (ms^2^)** |
| --- | --- | --- |
| Baratta L. et al. 2010 [2] | 51.8 ± 11 | 40.6 ± 18.5 |
| Frokjaer V. G. et al. 2006* [4] | 514.7 ± 961.9 | 25.5 ± 21.4 |
| Iga A. et al. 2003ˠ [5] | 141 ± 58 | 28 ± 36 |
| Ko F.Y. et al. 2013** [6] | 706.3 ± 2.4 | 135.6 ± 5 |
| Lazzeri C. et al. 1997 ˠ [7] | 15 ± 1 | 20 ± 1 |
| Mani A.R. et al. 2008 [8] | 188 ± 253 | 26 ± 34 |
| Milovanovic B. et al. 2009** [9] | 1096.6 ± 2.7 | 333.6±3.2 |
| Miyajima H. et al. 2001 [10] | 177.5 ± 94.0 | 77.2 ± 57.3 |
| Moller S. et al. 2012* [11] | 473.8 ± 514.3 | 151.3 ± 252.8 |

**= Reported as mean and interquartile range; **= Reported as natural log; ˠ= Reported as day and night [day recorded]. All other indices are reported as mean ± standard deviation.*

*HF, High Frequency; SD, Standard Deviation; ms, Millisecond.*

**Table S10 –** Reported Low Frequency (LF) of included studies.

| **Author** | **Normal LF (Mean±SD) (ms^2^)** | **Abnormal LF (Mean±SD) (ms^2^)** |
| --- | --- | --- |
| Frokjaer V. G. et al. 2006* [4] | 1171.1 ± 1726.2 | 74.5 ± 51.9 |
| Iga A. et al. 2003ˠ [5] | 213 ± 51 | 33 ± 23 |
| Ko F.Y. et al. 2013** [6] | 1085.7 ± 2.3 | 183.1 ± 4.4 |
| Lazzeri C. et al. 1997 ˠ [7] | 82 ± 2 | 79 ± 1 |
| Mani A.R. et al. 2008 [8] | 191 ± 145 | 29 ± 51 |
| Milovanovic B. et al. 2009** [9] | 3604.7 ± 2.7 | 943.9 ± 2.6 |
| Moller S. et al. 2012* [11] | 647.5 ± 865.2 | 150.3 ± 174.6 |

**= Reported as mean and interquartile range; **= Reported as natural log; ˠ= Reported as day and night [day recorded].*

*LF, Low Frequency; SD, Standard Deviation; ms, Millisecond.*

**Table S11 –** Reported Very Low Frequency (VLF) of included studies.

| **Author** | **Normal VLF (Mean±SD) (ms^2^)** | **Abnormal VLF (Mean±SD) (ms^2^)** |
| --- | --- | --- |
| Frokjaer V. G. et al. 2006* [4] | 734.6 ± 932.2* | 103.9 ± 71.7* |
| Negru R.D. et al. 2015 [13] | 2045.96 ± 1311.92 | 1385.86 ± 840.08 |

**= Reported as mean and interquartile range.*

*VLF, Very Low Frequency; SD, Standard Deviation; ms, Millisecond.*

**Table S12 –** Reported Low Frequency/High Frequency Ration (LF:HF) of included studies.

| **Author** | **Normal LF:HF (Mean±SD) (ms^2^)** | **Abnormal LF:HF (Mean±SD) (ms^2^)** |
| --- | --- | --- |
| Iga A. et al. 2003ˠ [5] | 1.14 ± 0.85 | 1.27 ± 0.22 |
| Miyajima H. et al. 2001 [10] | 1.20 ± 1.01 | 3.56 ± 2.31 |
| Moller S. et al. 2012* [11] | 1.27 ± 0.32 | 0.62 ± 0.15 |

**=Reported as mean and interquartile range; ˠ=Reported as day and night (day recorded).*

*LF:HF, Ratio of Low Frequency/High Frequency; SD, Standard Deviation; ms, Millisecond.*

**Table S13 –** Reported HRV non-linear indices; SD1, SD2, Sample Entropy (SampEn) and Detrended Fluctuation Analysis- (DFA α1) of included studies.

| **Author** | **Normal SD1 (Mean±SD)** | **Abnormal SD1 (Mean±SD)** | **Normal SD2 (Mean±SD)** | **Abnormal SD2 (Mean±SD)** | **Normal SampEn (Mean±SD)** | **Abnormal SampEn (Mean±SD)** | **Normal DFA α1 (Mean±SD)** | **Abnormal DFA α1 (Mean±SD)** |
| --- | --- | --- | --- | --- | --- | --- | --- | --- |
| Ko F.Y. et al. 2013 [6] | NS | NS | NS | NS | NS | NS | 1.14 ± 0.15 | 0.89 ± 0.25 |
| Mani A.R. et al. 2008 [8] | 23.2 ± 14.0 | 8.8 ± 5.9 | 57.8 ± 18.4 | 23.3 ± 12.6 | 2.89 ± 0.52 | 2.05 ± 0.60 | NS | NS |

*SD1, Poincare plot Standard Deviation perpendicular to the line of identity; SD2, Poincare plot Standard Deviation along the line of identity; SampEn, Sample Entropy; DFA α1, Short-term fluctuation of Detrended Fluctuation Analysis; NS, Not Shown.*

Database: Embase Classic+Embase <1947 to 2020 February 19>

Search Strategy:

--------------------------------------------------------------------------------

1	heart rate variability/ or autonomic nervous system/ (95567)

2	Heart Rate Varia*.mp. [mp=title, abstract, heading word, drug trade name, original title, device manufacturer, drug manufacturer, device trade name, keyword, floating subheading word, candidate term word] (34850)

3	liver disease/ or liver cirrhosis/ or chronic liver disease/ (254250)

4	1 or 2 (101756)

5	3 and 4 (250)

6	limit 5 to conference abstract status (58)

7	5 not 6 (192)

8	limit 7 to english language (154)

9	limit 8 to editorial (8)

10	8 not 9 (146)

11	limit 10 to "reviews (best balance of sensitivity and specificity)" (24)

12	10 not 11 (122)

**Figure S1 (a).** Search strategy for Embase.

Database: Ovid MEDLINE(R) and Epub Ahead of Print, In-Process & Other Non-Indexed Citations and Daily <1946 to February 20, 2020>

Search Strategy:

--------------------------------------------------------------------------------

1	Autonomic Nervous System/ (26244)

2	Heart Rate Varia*.mp. [mp=title, abstract, original title, name of substance word, subject heading word, floating sub-heading word, keyword heading word, organism supplementary concept word, protocol supplementary concept word, rare disease supplementary concept word, unique identifier, synonyms] (18846)

3	Liver Diseases/ or Liver Cirrhosis/ (133838)

4	1 or 2 (40775)

5	3 and 4 (78)

6	limit 5 to (abstracts and "reviews (best balance of sensitivity and specificity)") (5)

7	5 not 6 (73)

8	limit 7 to english language (57)

9	limit 8 to editorial (0)

10	8 not 9 (57)

**Figure S1 (b).** Search strategy for Medline.

| Recent queries in pubmed | |  |  |
| --- | --- | --- | --- |
| Search | Query | Items found | Time |
| #6 | Search (((((((Liver Disease*) OR Chronic Liver Disease*) OR Liver Cirrhosis) OR Cirrhotic*)) AND ((heart rate variability) OR HRV))) NOT (((((((Liver Disease*) OR Chronic Liver Disease*) OR Liver Cirrhosis) OR Cirrhotic*)) AND ((heart rate variability) OR HRV)) AND Review[ptyp]) Filters: Humans Sort by: Author | 66 | 09:27:41 |
| #5 | Search (((((((Liver Disease*) OR Chronic Liver Disease*) OR Liver Cirrhosis) OR Cirrhotic*)) AND ((heart rate variability) OR HRV))) NOT (((((((Liver Disease*) OR Chronic Liver Disease*) OR Liver Cirrhosis) OR Cirrhotic*)) AND ((heart rate variability) OR HRV)) AND Review[ptyp]) | 79 | 09:22:50 |
| #4 | Search (((((Liver Disease*) OR Chronic Liver Disease*) OR Liver Cirrhosis) OR Cirrhotic*)) AND ((heart rate variability) OR HRV) Filters: Review Sort by: Author | 6 | 09:21:20 |
| #3 | Search (((((Liver Disease*) OR Chronic Liver Disease*) OR Liver Cirrhosis) OR Cirrhotic*)) AND ((heart rate variability) OR HRV) | 85 | 09:21:15 |
| #2 | Search (((Liver Disease*) OR Chronic Liver Disease*) OR Liver Cirrhosis) OR Cirrhotic* | 249446 | 09:19:20 |
| #1 | Search (heart rate variability) OR HRV | 27650 | 09:17:44 |

**Figure S1 (c).** Search strategy for PubMed.

**Figure S1 (a-c).** Search strategies for Embase (a), Medline (b) and Pubmed (c) databases.

**Figure S1 (a-c).** Search strategies for Embase (a), Medline (b) and Pubmed (c) databases.
